# Consequences of Mismatch, Misalignment and Rotation of Toric Intraocular Lenses in Refractive Cataract Surgery Part 1. It Ain’t 30. The True Angle of Doom

**DOI:** 10.1101/2020.06.09.20126987

**Authors:** Samir I Sayegh

## Abstract

**Purpose:** To demonstrate that the total loss of astigmatism as a consequence of misalignment or rotation of a toric intraocular lens (tIOL) can occur much earlier than the widely believed and taught 30 degrees. To give a precise surgically useful estimate of that value. To clarify the role of mismatch and misalignment of toric intraocular lenses in cataract surgery beyond what is commonly recognized in the literature and make corresponding surgical recommendations.

**Setting:** Private Practice and Research Center. The EYE Center. Champaign, IL, USA.

**Design:** Formal Analytical Study

**Methods:** The astigmatism addition approach is used in its simplest form along with analytical tools to derive new results concerning mismatch, misalignment and rotation of toric intraocular lenses.

**Results:** The often stated results of total loss of astigmatic correction by 30-degree rotation and 3.3 % loss per degree represent a usually poor approximation to realistic surgical cases. We show how they constitute a very special case in the context of a more general framework relevant to procedures performed by refractive cataract surgeons dealing with the surgical correction of astigmatism with tIOLs. Total loss of astigmatic correction can occur with as little as 20 degrees of misalignment and less than 10 degrees of tIOL rotation. A practical approximation for that *angle of doom*, Δ, in the surgically relevant range can be expressed by **Δ** ≈ **30** − **15 *ω*** degrees, where 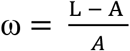 is the fractional *overcorrection* of L, the cylinder of the tIOL, and A, the astigmatism to be corrected. Similarly for undercorrection we show that **Δ** ≈ 3**0** + **15 *u*** degrees where 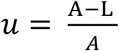 represents the corresponding fractional undercorrection. That is to say the angle of doom is extended beyond the 30 degrees for cases of undercorrection of the astigmatism. We also demonstrate that overcorrection of astigmatism results in a significantly faster decline in astigmatism correction per degree of misalignment/rotation. The significant clinical implications and surgical recommendations, including for optimal degree of overcorrection, are a natural consequence of these novel results.

**Conclusions:** Total loss of astigmatism correction can occur at a significantly smaller angle than commonly believed and overcorrected astigmatism residual rises with tIOL misalignment or rotation significantly faster than undercorrected astigmatism. We provide the methodology and explicit solution for determining this behavior.

## INTRODUCTION

As many as half of patients presenting for cataract surgery can potentially benefit from simultaneous correction of astigmatism, present initially and possibly induced by the procedure^1^. Surgical methods of correcting astigmatism have included limbal relaxing incisions (LRI), corneal incisions and toric intraocular lenses (tIOL). tIOLs have several advantages and have been used increasingly in the past decade. One potential limitation of tIOLs however concerns possible rotation and more generally misalignment due to rotation but also to positioning and initial inaccuracies in the estimate of the cross cylinder astigmatism to be corrected after the incisions have been performed and the procedure completed. Several investigators have looked at the consequences of tIOL rotation or misalignment both theoretically and in actual clinical studies. The occurrence of such tIOL rotations has also been documented by direct and indirect methods since the inception of the field to the present. The significant popularity of an online calculator to analyze the possibility of improving a refractive toric surprise is an indication that in clinical practice the occurrence of such surprises is common^2^. *The most common statement concerning the rotation or misalignment of a tIOL is that it loses 100% of its astigmatic correction effect at 30 degrees of misalignment at the rate of 3*.*3% per degree*, getting worse afterwards with the residual astigmatism exceeding the initial astigmatism to be corrected^3,4,5,6,7,8,9,10,11,12,7,13,14,15^. When this statement is made, the underlying assumption is that the magnitude of the cylinder of the tIOL is equal to the magnitude of the astigmatism to be corrected. *In surgical practice, the fulfillment of this assumption is neither common nor realistic*. One reason is that most tIOLs used in practice have a discrete set of cylinder values. For example, T3, T4, correcting about 1 diopter (D) or 1.5 D of astigmatism at the corneal plane, respectively but nothing in between. Even if the astigmatism to be corrected at the corneal plane were perfectly known, say 1.27 D, this would not match the cylinder of either T3 or T4 tIOLs and the usual matching assumption would be immediately violated resulting in undercorrection or overcorrection depending on which tIOL is chosen. A second reason, also relating to the tIOL, is the fact that the ideal toricity ratio may not be used because of limitations of the toric calculator or uncertainty in the effective lens position (ELP), a variable contributing to the toricity ratio and thus to the value of the tIOL cylinder at the corneal plane^16^. Additional reasons, relating to the astigmatism A to be corrected at the corneal plane, include uncertainties on measurements of the magnitude and meridian of its dominant anterior contribution^17,18^, uncertainties on the posterior astigmatism, as well as uncertainty on the values of surgical induced astigmatism, with estimates of SIA ranging from zero to half a diopter for similar clear corneal incisions^19,12^. All these reasons contribute to a high likelihood of some degree of mismatch between the tIOL cylinder and the estimated astigmatism to be corrected, including different tIOL recommendations by different calculators of the same manufacturer^20^. This is confirmed by the fact that even after perfect (re)alignment, a residual astigmatism of up to or beyond 0.75 D, often occurs, and up to 0.75 D is deemed acceptable^221^.

So the question “At what angle of misalignment of the tIOL is the astigmatic correction nullified?” needs a more sophisticated answer. We designate this critical angle of misalignment as the “*angle of doom*” in analogy to the “triangle of doom,” a concept from general surgery suggesting an area to be avoided during hernia repair^22^. Since the inception of the field of tIOLs the common belief has propagated that this angle is equal to 30 degrees. We show that this is a very special case that rarely corresponds to clinical reality. Literature dating back to the 19^th^ and 20^th^ century establishes the basic method of adding astigmatism and suggests that overcorrection induces more rotational residual astigmatic error^23,24,25,26,27,28,29,30^, which implies that the nullification of an overcorrected astigmatism is reached at an angle smaller than 30 degrees. The concept does not appear to have filtered to modern day refractive cataract surgery with tIOLs with few notable exceptions usually limited to numerical examples^29,31,30,32^. A straightforward, full and accurate development of this theme is presented here. In a companion paper we give a rigorous derivation of the optimal degree of overcorrection to be accepted when selecting a tIOL. The corresponding recommendations are somewhat different than those adopted by major tIOL manufacturers or those made recently in the surgical literature^33^.

## MATERIALS AND METHODS

In this study we use simple geometric means to represent the effect and loss of effect of astigmatism corrected via a toric intraocular lens. We also use the simplest possible nomenclatures and representation. To the extent possible we use a single letter to represent a measurement or a physical quantity. Since we will be dealing with the presumed corneal astigmatism to be corrected A, the astigmatism of the correcting tIOL at the corneal plane, L and the residual astigmatism R, we chose a triangle representation where L,R and A correspond to the length of the three sides, where the angle 2λ between L and A in the astigmatism “vector” representation is double the angle λ between L and A as measured in the eye itself. This “double angle” representation, relevant to differential geometry, optometry and ophthalmology, goes back at least to the 18^th^ and 19^th^ century^23,24,25^ and has been reintroduced to modern refractive cataract surgery in the past decades. With A= |A|, L = |L| the magnitudes of the “vectors” A and L, will always be considered positive and R_0_ = |A-L| is the magnitude of the mismatch between A and L.

We consider 6 cases as illustrated in Figure 1

1. Perfect alignment and magnitude match. λ=0, L = A. R = R_0_ = 0
2. Perfect alignment with undercorrection. λ=0, L < A. R = R_0_
3. Perfect alignment with overcorrection. λ=0, L > A. R = R_0_
4. Misalignment with magnitude match. λ ≠ 0. L = A
5. Misalignment with undercorrection. λ ≠ 0. L < A
6. Misalignment with overcorrection. λ ≠ 0. L > A

We do not distinguish between the cases where misalignment is clockwise or counterclockwise as the results are essentially equivalent for the purpose of this analysis. Figure 1 represents the 6 cases as listed.

**Figure 1.**
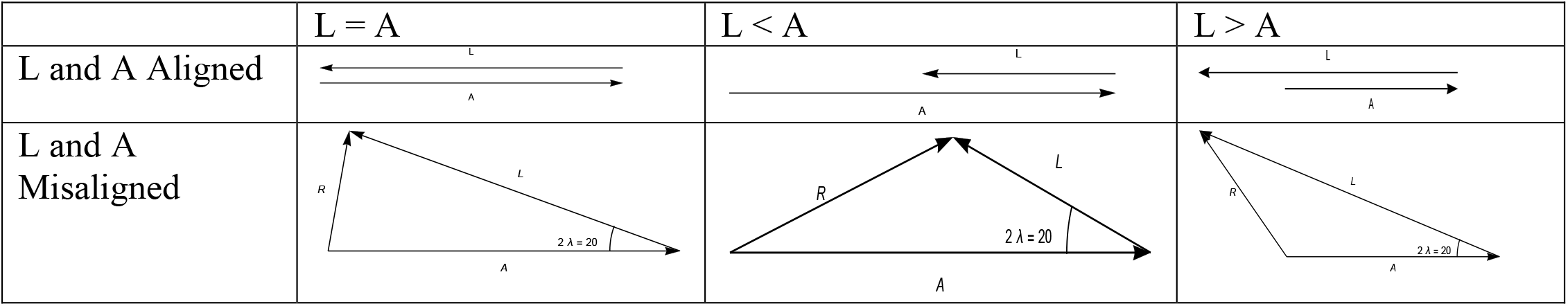
Six cases illustrating all possibilities of mismatch and misalignment of astigmatism A to be corrected by tIOL cylinder L, both at the corneal plane.

We designate the value of the angle λ for which R becomes equal to A by Δ, *the angle of doom*, and we start by illustrating five important points later developed in this paper (Figure 2):

1. Matched but misaligned L and A lead to R = A at Δ=30 degrees.
2. Mismatched and misaligned L and A give rise to R=A at Δ > 30 degrees for undercorrection.
3. Mismatched and misaligned L and A give rise to R=A at Δ < 30 degrees for overcorrection.
4. The departure of the value of Δ from 30 degrees can be very significant.
5. The maximum value of Δ is 45 degrees.

**Figure 2:**
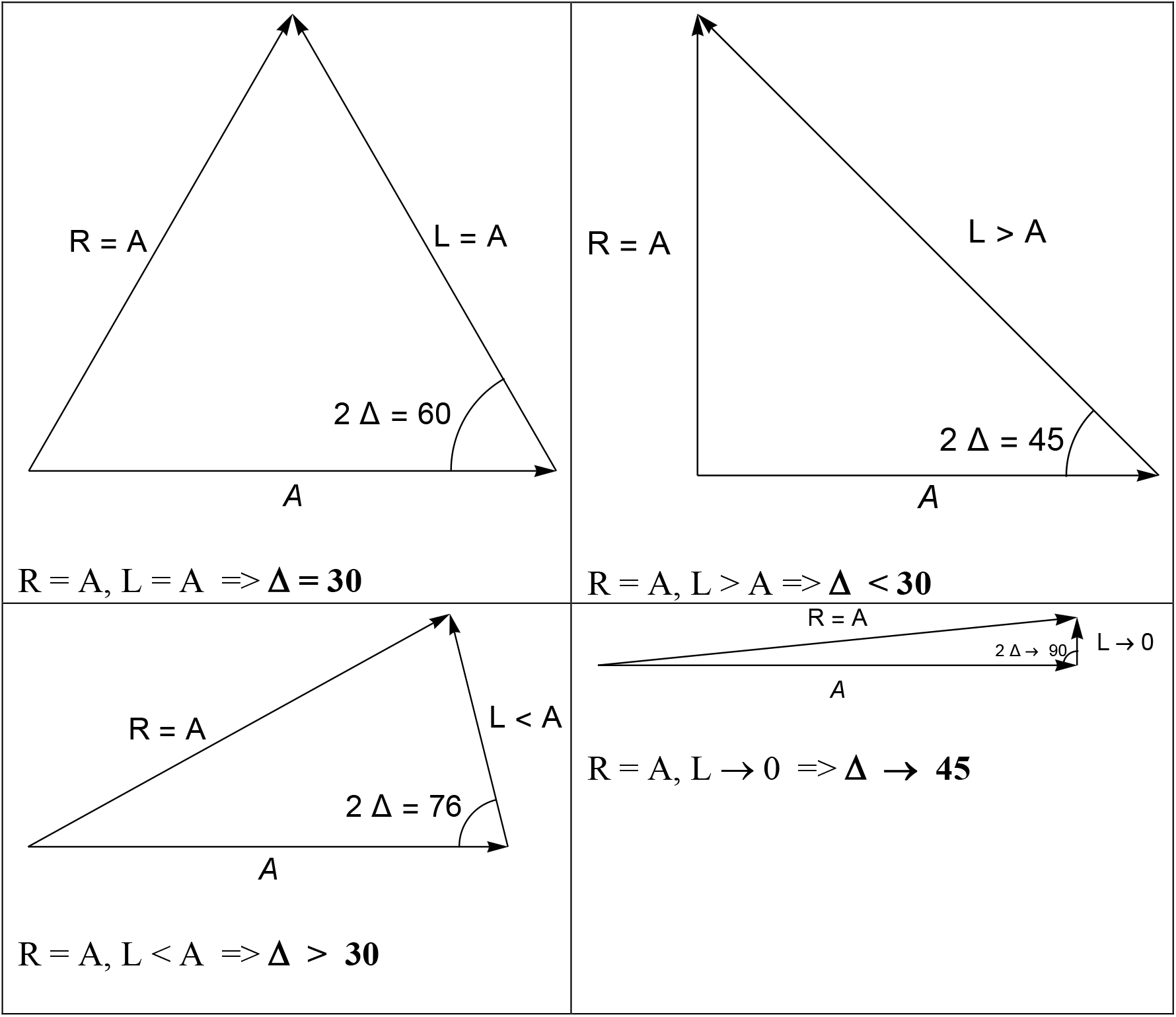
Illustrates the cases of 1) perfect match of A and L resulting in an equilateral triangle and thus 2 Δ = 60 degrees and Δ = 30 degrees, the “classical” result. 2) Overcorrection L ≃ 1.4 A and Δ = 22.5 and 3) Undercorrection L = 0.5 A and λ ≃ 38 degrees. 4) Limit of “barely toric” IOL with Δ → 45 degrees.

Consider the case L=A and set R=A. We now have L=R=A, and thus an equilateral triangle with all three angles equal and thus 2Δ = 60 degrees. Δ = 30 degrees. This is the case most commonly cited in the literature.

For the case L > A we can rotate L until its tip lays just above the “tail” of A (i.e. has the same horizontal coordinate) the triangle formed by LRA is now a right angle triangle, but the condition R=A also makes it isosceles. The two non-right angles are thus each equal to 45 degrees. So 2 Δ = 45 and Δ = 22.5 degrees. The length of L can be obtained from the Pythagorean theorem and is simply √2 A ∼ 1.4 A. This illustrates that *an overcorrection of about 40% will result in a drop of 25% in* Δ, and that this angle is actually much closer to 20 than it is to 30 degrees. This presents us with a less tenable surgical scenario, especially if other uncertainties contribute to potential misalignment.

We now illustrate the case of undercorrection. We use L= ½ A and rotate till R=A. The isosceles triangle with base L = ½ A has angle 2Δ ≅ 76 degrees and thus Δ ≅ 38 degrees. This demonstrates on an easily visualized example that undercorrection appears to increase the angle of doom. The last triangle case we observe is that of a very thin triangle as L becomes very small and tends to zero. The two equal angles of the isosceles triangle required for the equality of A and L remain equal in the limit and now add up to 180 degrees. We thus have 2 Δ = 90 and Δ = 45 degrees. This illustrates the limiting value of Δ.

### General Case

We now analyze the general case using simple trigonometry and elementary algebraic manipulations. A more geometric approach is deferred to a review article.

Applying the law of cosines to the ALR triangle of Figure 3, we obtain

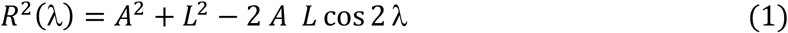

This can be rewritten as

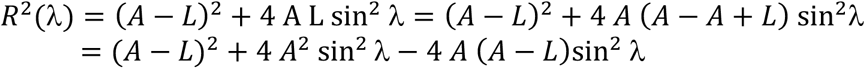

and eliminating L in favor of R(0) = R_0_ = |A-L|, we can rewrite

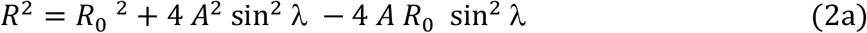

for undercorrection L < A, and

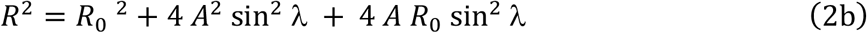

for overcorrection L > A

Note the similarities and differences between these two equations^1^.

**Figure 3.**
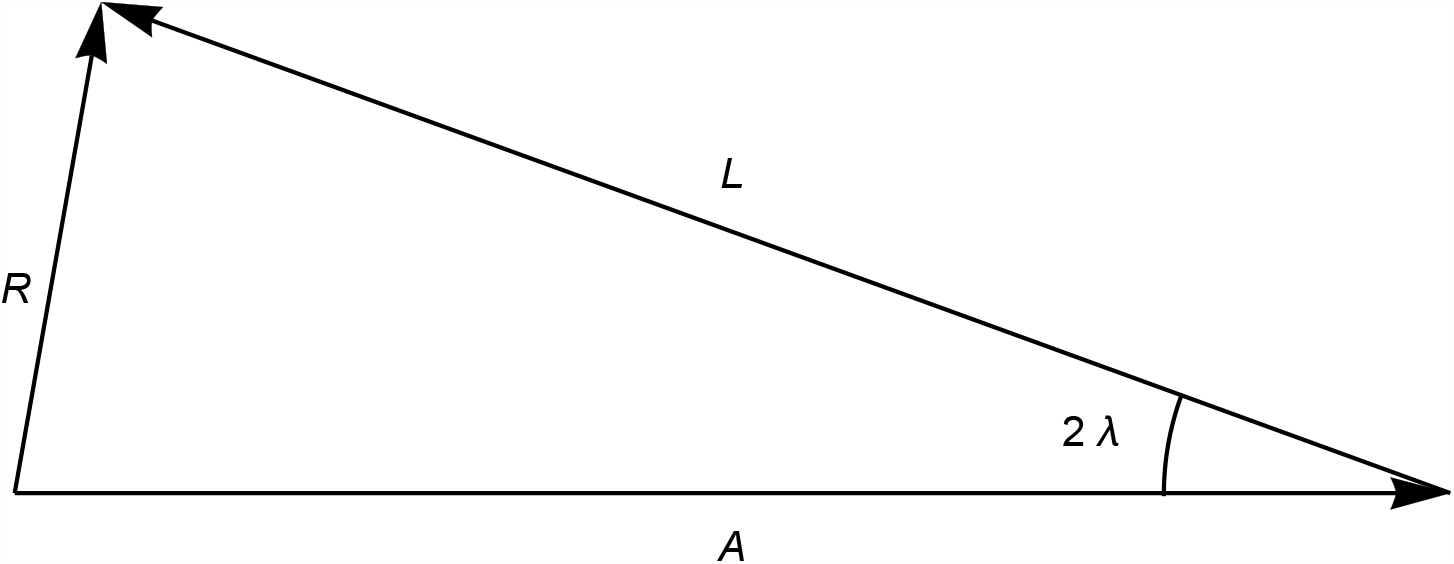
A triangle representation of astigmatism A with an attempted correction L that has rotated by an angle λ with respect to the intended meridian, with a residual astigmatism R. The geometric representation and residual calculation are based on a triangle with an angle 2λ between the sides, A and L. From elementary geometry an “SAS” (side angle side) triangle is uniquely determined and therefore both the residual astigmatism and its meridian are easily computable.

We take two additional steps to simplify the analysis. We introduce quantities defined relative to the astigmatism A to be corrected. Setting 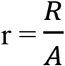 results in

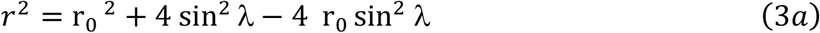

with 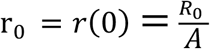
representing the fractional mismatch of L and A in the case of undercorrection. Similarly we have

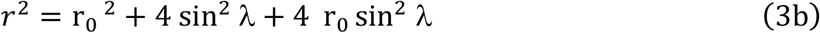

for the case of overcorrection.

This representation establishes that the relevant functional variables are ratios. The geometrical equivalent is considering one triangle, with one side having length = 1, to represent all similar triangles. All “actual” quantities can then be recovered by multiplying by the scaling variable (here A) at the end of the computation. This is the same as a change of unit where all quantities representing optical power are expressed in units of *A-diopter* instead of *diopter*, with the conversion factor being A.

We also define the astigmatism having been corrected S = A - R (so if the residual is zero for example then we have S = A and the full astigmatism has been corrected), and the fractional (or percentage) astigmatism having been corrected *s*,

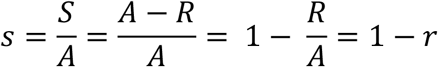

with

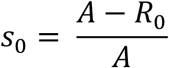

Clearly

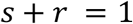

and

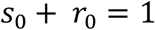

Any of the four representations, R, S, r and s, can then be chosen to graphically display the behavior of residual astigmatism as a function of mismatch and/or angle of misalignment: a) The residual error in diopters as a function of the increasing angle of misalignment, R(*λ*), typically an increasing function of *λ*, b) a residual error normalized to the value of A in diopters, r(*λ*), and where values are thus fractional, also an increasing function, c) The amount of astigmatism having been corrected S(*λ*) = A - R a decreasing function of *λ* and d) the fractional correction of astigmatism being corrected that is also a decreasing curve as a function of increasing angle *λ, s* (*λ*).

Figure 4 illustrates how one translates one representation to another by elementary algebraic operations. Results can be illustrated in one or more of the graphs of R, S = A - R, r or s, as a function of *λ* and we will usually indicate which representation is chosen and if we are in presence of under or overcorrection, unless the context makes it clear. The four options are illustrated in Figure 5 for the case of matching L and A.

**Figure 4.**
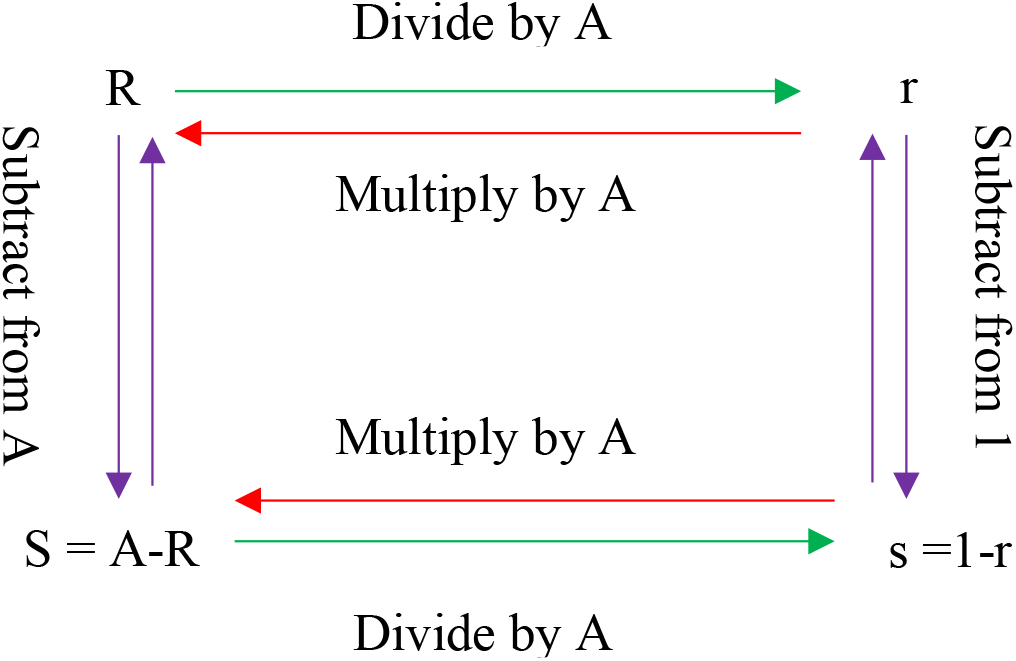
Navigating between 4 equivalent representations of residual astigmatism, 1) the full residual, R, 2) the fractional residual r, with respect to A, astigmatism to be corrected, 3) the magnitude of astigmatism having been corrected, S and 4) the fractional correction, s, with respect to A.

**Figure 5.**
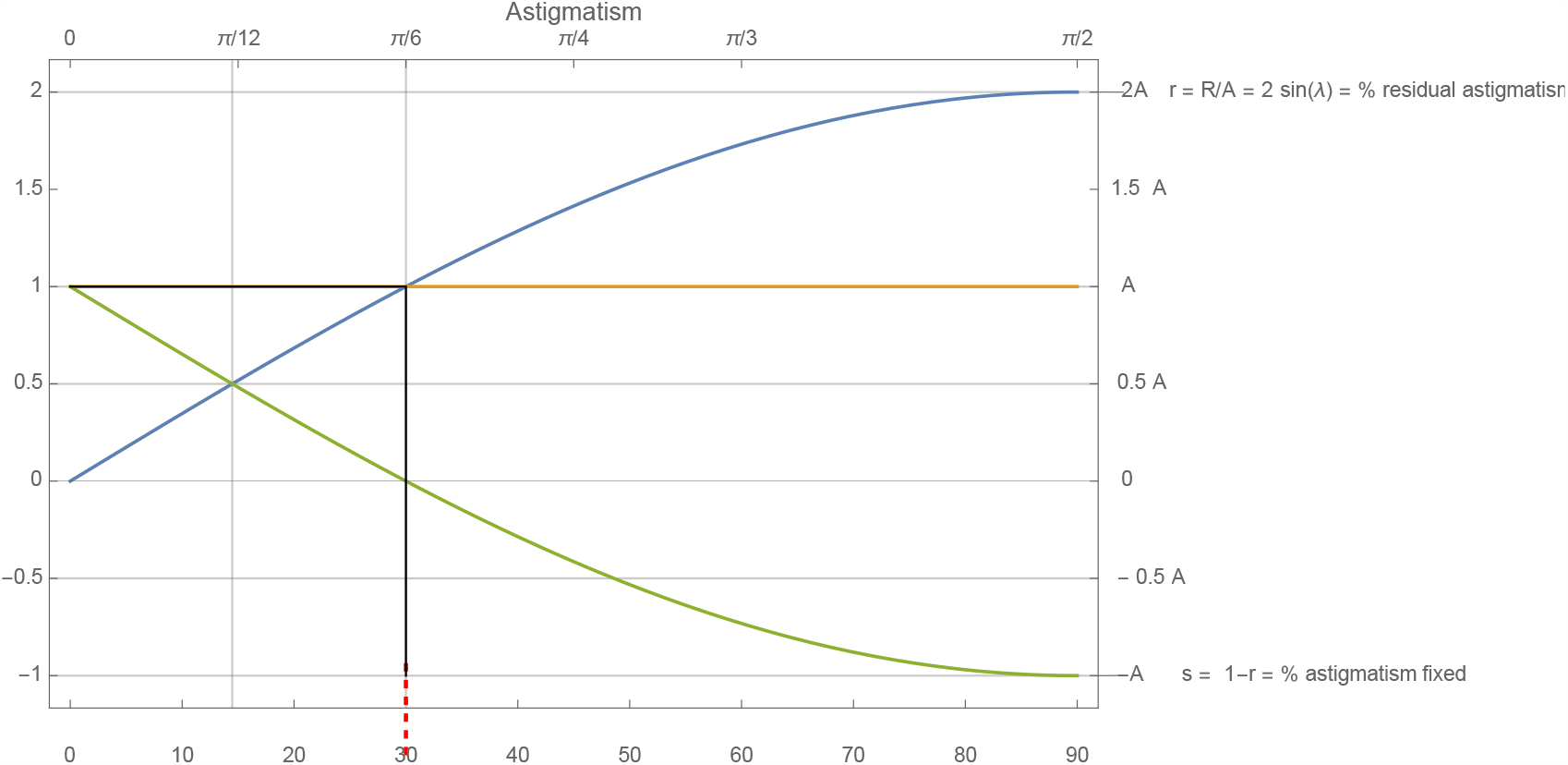
An astigmatism of cylinder value = A diopters, being corrected by a tIOL of L=A diopters as it rotates, results in residual astigmatism represented as magnitude of residual astigmatism (ascending curve), astigmatism corrected (descending curve) as full value of astigmatism (right scale) or its fractional representation wrt A (left scale).

A normalized increasing residual curve was given computationally for the matched case by Sanders, Grabow and Sheperd^3^ in 1992, with no functional dependence given explicitly but a sine dependence hinted at. That interesting chapter entitled “The Toric IOL” that ushered in the manufacturing and FDA approval of toric IOLs in the US dealt with a *non toric* STAAR IOL that was marked to examine rotational stability and simulate what would happen once tIOLs became available. In 2002, Till et al ^5^ presented a percent correction of astigmatism as a function of angle of rotation, also restricted to the matched case, and also numerically selecting or highlighting discrete values every 10 degrees and connecting them, with no mention of a specific functional dependence. Felipe et al. presented a normalized increasing astigmatism curve for the matched case and gave a functional dependence in the context of a discussion of matrix methods^29^.

## RESULTS

The expressions 2a and 2b can add significant insight to our understanding of tIOL astigmatism mismatch and misalignment. In particular, we observe the following

1. In the absence of misalignment, (*λ* = 0), we obtain the expected R_0_ term, in all cases of mismatch.
2. In the absence of mismatch (L = A, R_0_ = 0) and the presence of misalignment, we obtain the expected behavior of R as the angle of the tIOL changes, namely

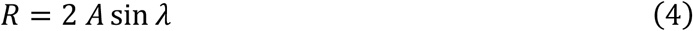

including when R=A (r =1) leading to *sin Δ = ½ and the often cited Δ* = 30 *degrees*.
3. In the presence of both mismatch and misalignment we obtain the sum of two terms corresponding to mismatch only (R_0_ but no *λ*) and misalignment only (*λ* but no R_0_), *in addition to a cross term that depends on both mismatch* R_0_ *and misalignment λ*.
4. The cross term has a different and *opposite sign contribution to the residual astigmatism in the case of undercorrection compared to the case of overcorrection*.
5. As a consequence of 4) and while the behavior of the residual astigmatism is symmetrical with respect to the side on which misalignment occurs, *it is not symmetrical with respect to the side on which mismatch occurs*.

We now turn our attention to the general case L ≠ A and compare it to the special case L = A. From Equations 3(a) and 3(b) we plot *r* as a function of angle λ of misalignment for cases of a perfect match of A to L (A = L) and two for a mismatch of 0.5 i.e(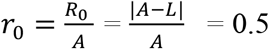) Figure 6. As an alternate representation, we also plot, *s*, the percent loss of astigmatism correction as a function of λ. As illustrated in Figure 7, these representations are equivalent but one or the other may be more appealing to the intuition of different readers.

**Figure 6.**
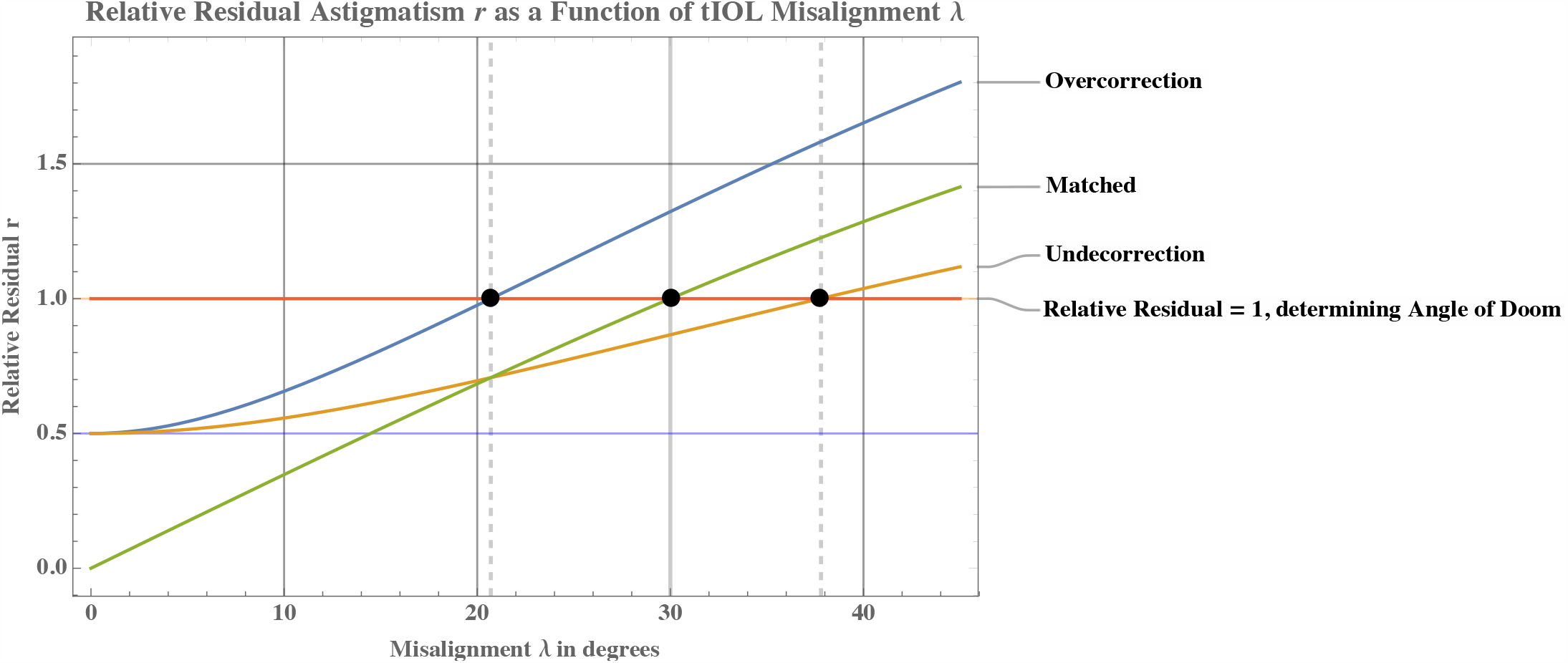
Relative residual error as a function of angle of misalignment for matched case (green) as well as over (blue) and undercorrection(orange). Angle of doom determined by intersection of each curve with r=1. Observe that for undercorrection Δ ∼ 40 degrees and for overcorrection Δ ∼ 20 degrees.

**Figure 7.**
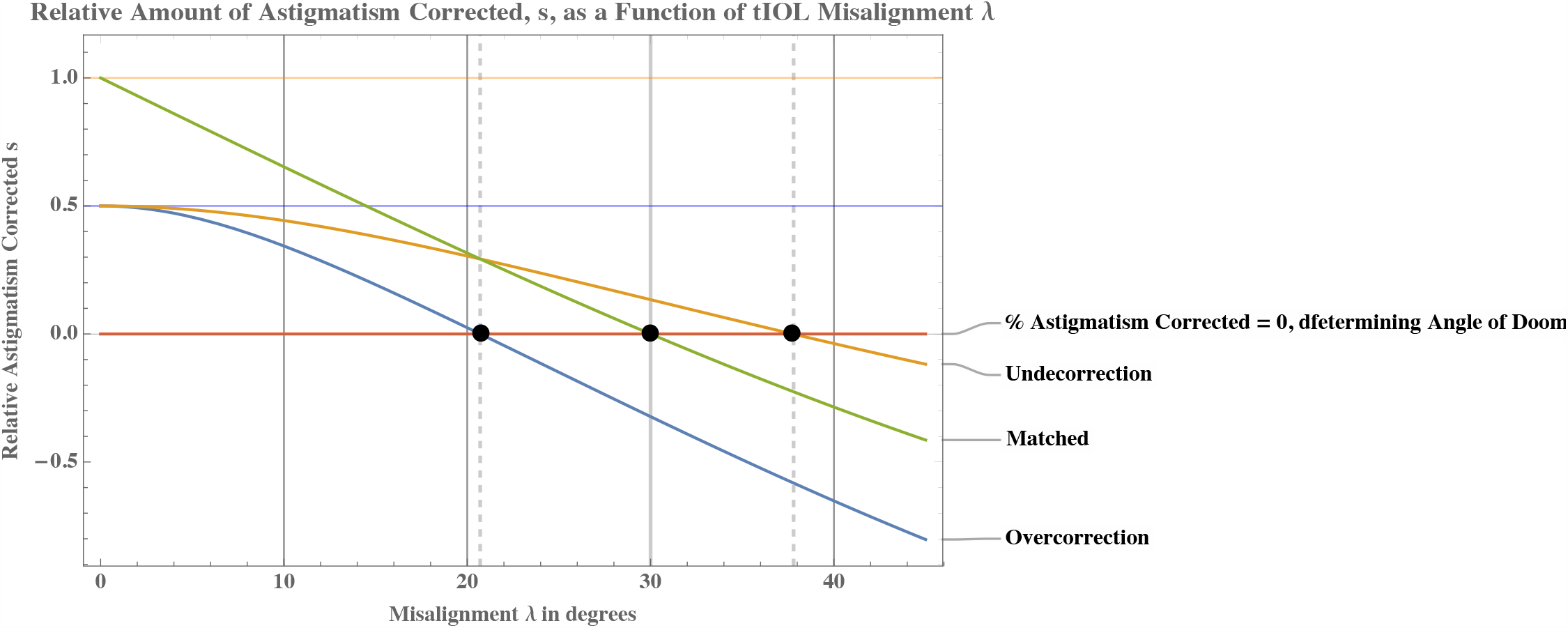
Relative amount of astigmatism corrected as a function of misalignment for matched case (green) as well as over (blue) and undercorrection(orange). Angle of doom determined by intersection of each curve with s=0. Clearly here too we observe that for undercorrection Δ ∼ 40 degrees and for overcorrection Δ ∼ 20 degrees.

The following observations can be made

1. In the case of perfect match, the rise of the residual *r* as a function of angle of misalignment is nearly linear. This corresponds to the nearly linear portion of the sine function^2^. In the *s* representation a corresponding early linear decay in % corrected astigmatism is seen.
2. For a perfect match, at 30 degrees, *r* = 1, meaning the residual is now equal to the original value of the astigmatism to be corrected. This is the known and often repeated result that captures this very special case but is far from representing most realistic surgical situations. In other words for the matched case, the angle of doom Δ = 30^°^.
3. The residual astigmatism in the *overcorrection* case L > A *rises faster* than it does for the undercorrection case L < A as the angle is rotated and the residual always remains larger. The angle of doom is thus reached earlier, and possibly much earlier, for the overcorrection case than for the matched or undercorrected case. For the specific 50% overcorrection or undercorrection displayed, we have Δ at just less than 21° in the case of overcorrection and Δ nearly 38° in the case of undercorrection. We can see that in quite realistic cases of overcorrection, the angle of doom can be about 10 degrees smaller than usually claimed. Similarly in cases of undercorrection, it can be about 10 degrees larger than the commonly taught value of 30 degrees.
4. The cases r_0_ = 0.5 and r_0_ = 0 actually cross, with the graph for undercorrection yielding less residual past the intersection point. Crossing occurs also around 21° in this case as indicated on the graph. This is an indication that for a certain degree of misalignment, *undercorrection may actually be preferred not only to overcorrection, but also to an exact match of the tIOL to the astigmatism to be corrected*. One can easily show that for r_0_ = 0.1, undercorrection becomes preferable to perfect match at about 9 degrees, for the case r_0_ = 0.2, at about 13 degrees and for r_0_ = 0.3 it occurs at about 16 degrees. A full discussion will be presented in a companion paper and its clinical and surgical consequences will be shown to be significant.

**Predicting the Angle of Doom: When do we totally lose the effect of astigmatic correction?**

The question is usually asked in terms of an angle of alignment and/or rotation of the tIOL relative to the *presumed* meridian of the astigmatism to be corrected. The answer clearly depends on L, and more specifically on 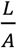

Returning to Equation (1) we now consider A and L having arbitrary values, not necessarily equal. We first establish the conditions for *R* = *A*. Recalling Equation (1)

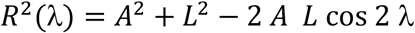

We divide by A^2^, define 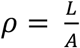 and recall that 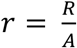, yielding

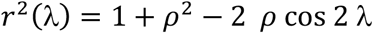

Requiring R = A, implies *r* = 1, and results in the condition for λ = Δ, the angle of doom,

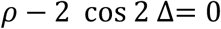

Or

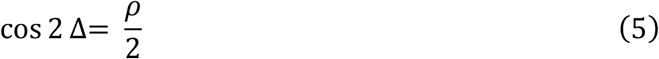

A similar condition, slightly less elegantly expressed, was given by Felipe et al^29^ in the context of matrix methods with few of its implications explored.

Here we propose to 1) explore a few *key values* before 2) giving a full expansion of the function and the subject surrounding it, by explicitly inverting Equation (5) to give the values of Δ as a fu*nction of ρ* and then 3) provide an excellent approximation of Δ, for both under and overcorrection, that can readily be used in *most clinical situations*.

For *ρ* = 0, the “non toric” case, we have 2 Δ = 90 degrees and Δ = 45 degrees, the proper limit presented geometrically in an earlier section (see Figure 2). The effect of a very low correcting tIOL will be totally lost at 45 degrees misalignment or less. For *ρ* = 1, the matched case, and the one usually discussed or assumed in much of the literature, we have 2 Δ = 60 degrees and thus Δ = 30 degrees, the usually quoted result. Finally for *ρ* = 2, ie overcorrection by the full initially present astigmatism, A, we have already reached “doom” with no need for misalignment and indeed cos 2 Δ= 1 yields Δ= 0, as expected.

One can solve Equation (5) for Δ and plot the result (Figure 8) as a function of *ρ* for 0 ≤ *ρ* ≤ 2

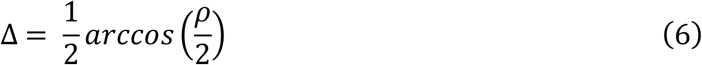

From Figure (8) one confirms the cancellation angle is 30 degrees for 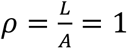 as shown but more interestingly that the value of Δ can be significantly larger for L< A and significantly smaller for L > A, as illustrated in specific examples in the introduction. Here again we can see that a 50% undercorrected eye can extend the angle Δ to almost 40 degrees while a 50% overcorrection can reduce Δ to about 20 degrees, as shown explicitly in Figure 6 and Figure 7.

**Figure 8.**
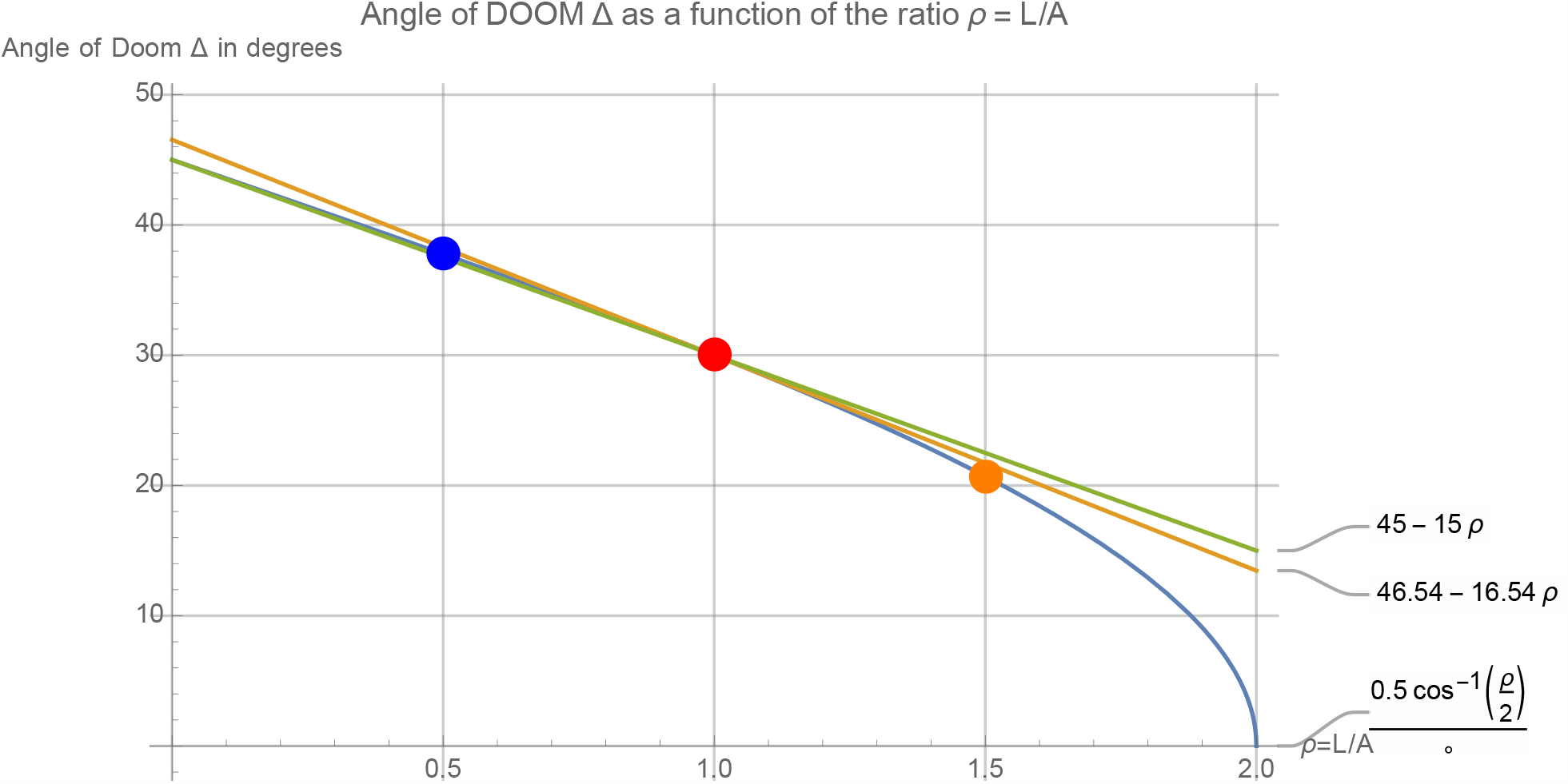
Angle of Doom Δ as a function of 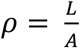 with linearization around (1,30)

The function in Equation (6) as seen on graph in Figure 8 can now be linearized by an expansion in a Taylor series around the value 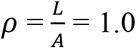 to yield Δ ≅ 46.54 - 16.54 *ρ*. This gives the correct value of 30 for *ρ* =1 and is a very good practical approximation for *ρ* in the range between zero and 1.5 and excellent in the clinically practical range of 0.5 to 1.5. The more practical ∼ 45 – 15 *ρ* has the additional advantage of matching the limit at *ρ* = 0 to its correct expected 45 degrees limit, while also predicting the 30 degrees of the “classic” *ρ* = 1 case. In any case the approximation is excellent in the full range of 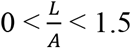 and it is clear that past a value of 1.5 the angle falls very rapidly to zero consistent with the fact that the correcting L value is nearing double the value of the astigmatism to be corrected. For clinical estimate in the range 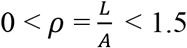 we thus adopt

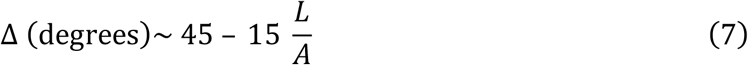

which can be rewritten

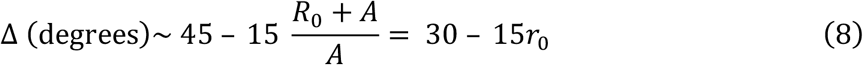

Or, for an overcorrection

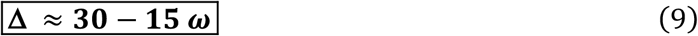

Similarly for undercorrection a change of variable yields

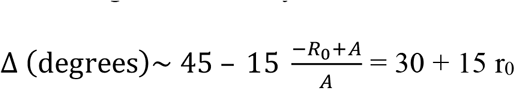

and we can express Δ for an undercorrection as

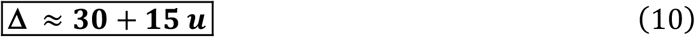

**These expressions for Δ are centered on 30 degrees, the value often cited, but make it clear that overcorrection would result in a significantly smaller angle of doom and conversely, undercorrection extends the range of misalignment for which some astigmatic correction is still beneficial**. The expressions are very practical and facilitate the immediate visualization of the effect and give an accurate estimate in the most relevant clinical range of 0 ≤ *L* ≤ 1.5 *A* covering a continuous range including both significant under and overcorrection. As pointed out previously they could be expressed in one equation with a sign indicating the undercorrecting or overcorrecting nature of the situation. For the sake of clarity of this and further discussions we prefer to continue to separately and explicitly display the sign dependence on over and undercorrection.

## DISCUSSION AND CONCLUSION

This work is part of a tradition of description, analysis and development of computational tools dealing with astigmatism in the human eye that extends back more than two centuries and, of its surgical correction with toric intraocular lenses that extends more than two decades. Improvement in analysis, surgical technique and quality of the tIOLs available have brought us much closer to the goal of simultaneous elimination of undesirable refractive error at the time of cataract surgery. However, a belief has persisted as to the exact quantitative nature of the effect of a misalignment and rotation of the tIOL and the angle, here designated as the “angle of doom, Δ”, where the amount of correction provided by the tIOL reduces to zero. This angle is consistently believed to be 30 degrees, even though some computational studies in the context of clinical refraction, and more recently surgical correction, have indicated it may be otherwise. Using the simplest tools for analyzing astigmatism we give an exact treatment that shows convincingly that a clear distinction must be made between under and over correction. Our analysis also results in an exact expression for Δ as well as a clinically relevant and convenient approximation centered on 30 degrees, the value commonly believed to be the correct value. This work and a companion paper should contribute to the establishment of a rational approach to the selection of tIOLs for the surgical correction of astigmatism in general and especially at the time of refractive cataract surgery.

## Data Availability

All data needed and not appearing in the manuscript is available by emailing the author

## Footnotes

1 An alternative approach is to define *R*_*0*_ as a signed quantity that is positive for undercorrection and negative for overcorrection resulting in one formula encompassing both under and overcorrection. For the remaining of the discussion we opt for the representation where *R*_*0*_ is always positive and we keep track separately of over and under correction. This is done for a number of reasons one of them being that a main point of the paper is to explicitly emphasize the distinction between under and overcorrection.

2 Because sin x ∼ x is a good approximation up to about 30 degrees, one can write 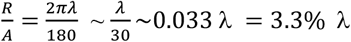 where λ is in degrees. These results (“Total loss of astigmatic correction at 30° and 3.3% loss per degree of rotation”) are often cited without clarifying either of the underlying assumptions, namely, magnitude match, L = A, and the linearization of the sine function. It is important to note that this approximation becomes increasingly poor beyond 30 degrees.

